# A Novel Multi-strain Vaginal Synbiotic is Effective in Optimizing the Vaginal Microbiome: Results from a Randomized, Placebo-controlled Clinical Trial

**DOI:** 10.1101/2024.05.20.24307554

**Authors:** Jacques Ravel, Eleni Greenwood Jaswa, Sara Gottfried, Miriam Greene, Susan Kellogg-Spadt, Dirk Gevers, Diane Harper

## Abstract

This research letter describes the results of a clinical trial of a novel vaginal synbiotic comprising three *L. crispatus* strains and a supportive nutrient complex. Results indicated that administration of the synbiotic led to an optimal vaginal microbiome dominated by *L. crispatus* (CST I) and additionally reduced the abundance of microbes associated with vaginal dysbiosis and the abundance of *Candida*, the most common source of vaginal yeast infections.

## Introduction

Compared to an optimal vaginal microbiome dominated by *L. crispatus* (CST I)^1^, vaginal dysbiosis is linked to several gynecological and obstetric conditions, including genital infections^2^ and pregnancy complications^3^. Biologically, vaginal dysbiosis is associated with increased vaginal inflammation and increased abundance of a diverse set of anaerobic microbes associated with degradation of the protective mucus barrier^4^. However, there are limited studies demonstrating that microbiome interventions with live beneficial organisms lead to an optimal vaginal microbiome. Accordingly, a novel vaginal synbiotic, comprising three *L. crispatus* strains sourced from stable vaginal microbiomes which maximized coverage of optimal *L. crispatus* genomic diversity plus a supportive nutritional complex, was evaluated in a clinical trial.

## Methods

Randomized, placebo-controlled clinical trial (n=70) with Part A comparing the multi-strain synbiotic vaginal tablet (VS-01) to placebo, and Part B comparing three formulations (vaginal VS-01 capsule, oral VS-01 capsule, over-the-counter oral supplement containing *L. crispatus, Lacticaseibacillus. rhamnosus, Lactobacillus gasseri*, and *Lactobacillus jensenii*). Participants were randomized to each arm for the intervention period following menses and assessed through Day 21 (D21) (**Supplement**). The primary outcome was safety, and the key secondary outcomes were the abundance of *L. crispatus* and taxonomic/functional microbiome profiles as assessed by metagenomic sequencing. Cytokine and chemokine profiles were established using a custom 33-plex Luminex panel.

## Results

We enrolled women of reproductive age (40% non-White, 60% White) between 18-54 years. Overall, all formulations were well-tolerated with a favorable safety profile. The multi-strain synbiotic vaginal tablet (VS-01) arm showed a significantly higher relative abundance of *L. crispatus* compared to placebo at D21 (p=0.024) among participants with baseline dysbiosis/non-optimal microbiome (**Figure 1A**). Consistent with the increase in *L. crispatus*, the VS-01 arm demonstrated a 90% conversion to an optimal CST I microbiome (defined as *L. crispatus* > 50%) compared to 11% in the placebo (p<0.002) among participants with baseline dysbiosis/non-optimal microbiome (**Figure 1B**). Further, the summed relative abundance of VS-01 strains increased significantly over baseline during the dosing regimen at D7 (p<0.05), D14 (p<0.001), and D21 (p<0.01). The vaginal tablet demonstrated superior conversion to an optimal CST I microbiome compared to vaginal capsule or oral formulations among participants with baseline dysbiosis/non-optimal microbiome (**Figure 1B**). The over-the-counter oral supplement did not lead to colonization.

**Figure 1.**
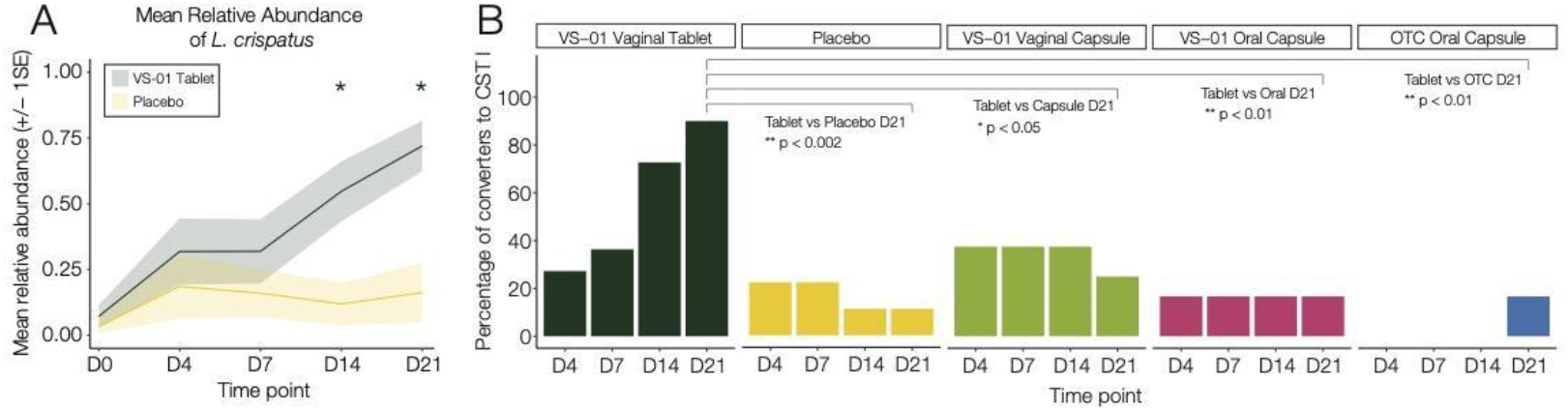
(A) Mean relative abundance of *L. crispatus* in the VS-01 vaginal tablet and placebo arms (n=19). * indicates p<0.05 difference between VS-01 tablet and placebo. (B) The proportion of participants who converted to CST I at each timepoint is shown for Part A and Part B among those with a baseline dysbiotic/non-optimal microbiome (n=39).

The vaginal VS-01 tablet reduced levels of microbes associated with vaginal dysbiosis which contributed to four key mechanistic findings (**Figure 2**). First, the mean relative abundance of *Candida* spp. was significantly reduced in the VS-01 arm (236-fold) compared to placebo (p=0.029) at D21 among participants with detectable *Candida* at baseline. Second, the relative abundance of *Gardnerella* spp. was significantly decreased in the VS-01 arm (p=0.013) on D21. These results aligned with preclinical data demonstrating that the strains comprising the synbiotic inhibited the growth of multiple strains of *Gardnerella* and *Candida* spp. using a parallel streak assay. Third, the abundance of mucin-degrading sialidase genes was significantly decreased in the VS-01 arm (p=0.036) at D21, suggesting the potential for improvement in the protective mucus barrier. Fourth, levels of the pro-inflammatory cytokine IL-1α were significantly decreased in the VS-01 arm at D21 (p=0.005).

**Figure 2.**
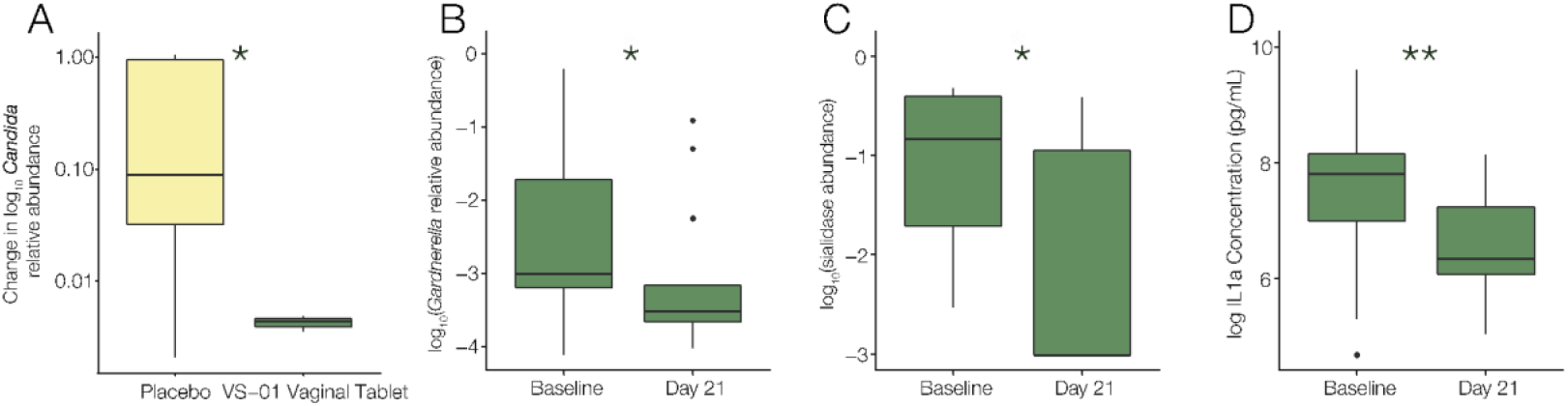
(A) VS-01 significantly decreased the mean abundance of *Candida* spp. compared to placebo from baseline to D21 (p=0.029). (B) VS-01 significantly decreased the abundance of *Gardnerella* spp., a key dysbiotic/non-optimal bacterium, from baseline to D21 (p=0.013). (C) VS-01 significantly decreased sialidase genes from baseline to D21 among participants with baseline detectable sialidase genes (p=0.036). (D) VS-01 significantly decreased the concentration of IL-1α from baseline to D21 (p=0.005).

## Discussion

This is the first study to demonstrate that a multi-strain vaginal synbiotic leads to an optimal CST I vaginal microbiome, a biomarker associated with a reduction in microbiome-mediated women’s health conditions. Biologically, the synbiotic reduced the abundance of microbes associated with vaginal dysbiosis and the abundance of *Candida*, the most common source of vaginal yeast infections. In summary, the multi-strain synbiotic effectively establishes an optimal and protective vaginal microenvironment.

## Data Availability

Selected data produced in the present study are available upon reasonable request to the authors.

## Acknowledgements

We thank Sheri Simmons PhD & Zain Kassam MD, MPH for contributions to data analysis/interpretation and review/revision of the manuscript; Gabriel A. Al-Ghalith PhD & Courtney Van Den Elzen PhD for contributions to data analysis; Michelle Davison PhD & Callahan Baker for contributions to clinical trial operations, and Raja Dhir for contributions on design concept.

## SUPPLEMENTAL METHODS

### Study Schematic

Part A of the trial (NCT05659745) assessed VS-01 compared to placebo, and Part B of the trial assessed several formulations (vaginal VS-01 capsule, oral VS-01 capsule, over-the-counter oral supplement). Participants collected baseline samples (D0) 5-10 days prior to the onset of menses (M). After the conclusion of menstrual bleeding, the intervention period was initiated (as shown in the schematic) and samples were collected prior to the next menses.

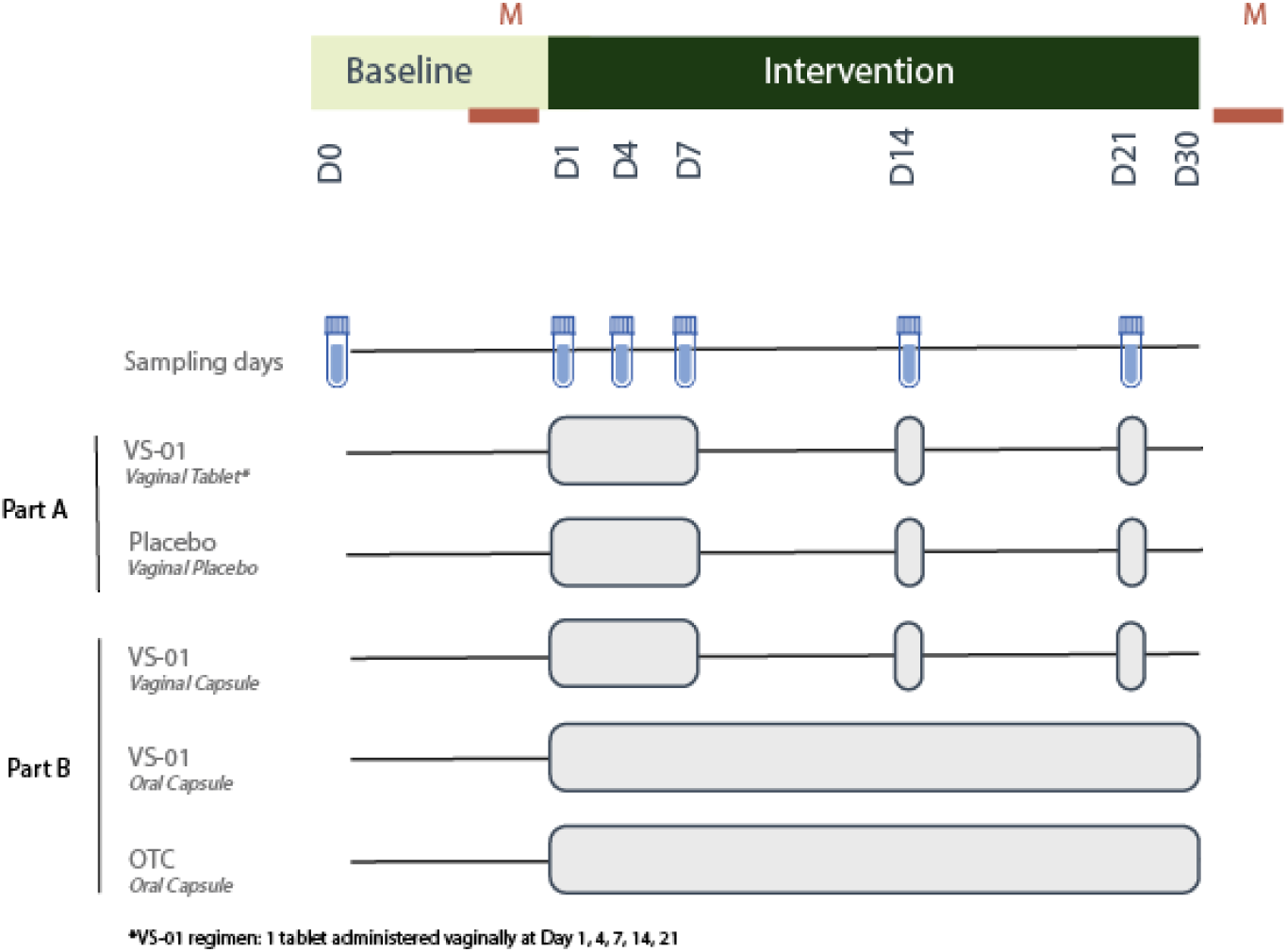

### Sample processing

Vaginal swabs were collected using the OMNIgene Vaginal kits (DNA Genotek, Ontario, Canada) which has been shown to be suitable for shipping at room temperature. Samples were shipped to Azenta Biosciences (Indianapolis, IL) for pre-processing with Proteinase K, aliquoting, and storage at -80°C as per OmniGene Vaginal kit recommendations. Aliquots were sent on dry ice to Zymo Research Corp. (Zymo Research, Irvine, CA) for DNA extraction, quantification, whole metagenomic library sequencing preparation and DNA sequencing. The ZymoBIOMICS-96 MagBead DNA Kit (Zymo Research, Irvine, CA) was used to perform DNA extraction according to the manufacturer’s instructions. 400μL of sample was used as input to the extraction.

Genomic DNA samples were profiled with shotgun metagenomic sequencing. Sequencing libraries were prepared with Illumina® DNA Library Prep Kit (Illumina, San Diego, CA) with up to 500 ng DNA input following the manufacturer’s protocol using 10 bp unique dual indices with Nextera® adapters (Illumina, San Diego, CA). Pooling and Post-Library QC: All libraries were pooled in equal abundance. The final pool was quantified using qPCR and TapeStation® (Agilent Technologies, Santa Clara, CA).

### Metagenomic Analysis

We developed a vaginal microbiome reference database using a combination of genomes from bacterial isolates and metagenome-associated-genomes (MAGs) assembled from public datasets. All quality-controlled samples were aligned to the vaginal microbiome database with XTree (0.92i), using the CAPITALIST algorithm [introduced in BURST (Al-Ghalith et al., 2020)] to normalize multi-mapping reads at the species level for species-level taxonomic characterization. XTree’s output included taxonomic counts, reference genome counts, and reference genome horizontal coverages (one table for proportion of the genome covered uniquely, and another for total proportion covered), for each sample. For all strain-level analyses, the unique and total genomic coverage proportions for the VS-01 strains were set to 0.88 (based on empirically maximizing variance in a test set) to conservatively demarcate the presence of these strains within metagenomic samples.

Fungal genomes were retrieved from NCBI RefSeq (R220) and taxonomy was assigned based on a previously established protocol (t2gg) (Al-Ghalith et al., 2020). An XTree database was constructed out of the downloaded fungal genome assemblies using the same parameters as the bacterial (compression level 2, k-mer size 29). Metagenomic samples were profiled using this database as described above for bacterial short reads Due to the sparsity of fungal reads compared to human and bacterial reads in most samples, a lenient total horizontal coverage threshold (0.1% coverage) was imposed to filter out only vanishingly low-abundance or spurious (contaminated) fungal genomes. Read counts were aggregated at the species level and reported when at least one genome within the species was at least 0.1% covered.

For identification and quantification of sialidase genes, the quality-controlled metagenomic reads were assembled using megahit v1.2.9 3, and gene inference was performed on the resulting contigs. The Kyoto Encyclopedia of Genes and Genomes’ (KEGG) V107 HMM database (Aramaki et al., 2020) was used to annotate each gene in each sample. The raw abundance of each gene was set to its contig’s vertical coverage in the sample, and normalized coverage was computed as the raw coverage divided by the total contig coverage in the sample, multiplied by 1 million (similar to TPM, but for gene abundances instead of transcripts).

### Cytokine Measurements

Vaginal swab samples were self-collected by participants and placed into 1 mL of a buffer that afford protein stability up to 7 days at room temperature. Samples were shipped at room temperature to Azenta and stored at -80 C until use. Vaginal swabs were thawed, and 100 μL of supernatant was aliquoted into 1.5 mL microfuge tubes and spun for 5 min at 12,000 x *g* to clarify.

Levels of a custom panel of 33 cytokines [epidermal growth factor (EGF), fibroblast growth factor (FGF), Eotaxin, transforming growth factor alpha (TGF-α), granulocyte colony stimulating factor (G-CSF), FMS-like tyrosine kinase 3 ligand (Flt-3L), granulocyte-macrophage colony-stimulating factor (GM-CSF), Fractalkine, interferon alpha (INFα), interferon gamma (INFγ), growth related protein (GRO), monocyte chemotactic protein 1 (MCP-1), interleukin 12 homodimer p40 (IL12p40), macrophage derived chemokine (MDC), interleukin 12 heterodimer p40 and p35 (IL12p70), platelet derived growth factor AA dimer (PDGFa), interleukin 13 (IL-13), platelet derived growth factor AB and BB dimer (PDGFab), soluble CD40 Ligand (sCD40L), interleukin 17 alpha (IL-17α), interleukin 1 receptor antagonist (IL-1RA), interleukin 1 alpha (IL-1α), interleukin 1 beta (IL-1β), interleukin 4 (IL-4), interleukin 6 (IL-6), interleukin 7 (IL-7), interleukin 8 (IL-8), interferon gamma-induced protein 10 (IP-10), macrophage Inflammatory protein 1 alpha (MIP-1α), macrophage inflammatory protein 1 beta (MIP-1β), RANTES (regulated on activation, normal T cell expressed and secreted) / Chemokine (C-C motif) ligand 5 (CCL5), tumor necrosis factor alpha (TNF-α), and vascular endothelial growth factor (VEGF)] were measured in the clarified swab extracts according to manufacturers’ instructions using a multiplexed immunoassay kit (MILLIPLEX®, MilliporeSigma Corp., Burlington, MA). A Bio-Plex 200 readout system was used (Bio-Rad, Hercules, California), which utilizes Luminex® xMAP fluorescent bead-based technology (Luminex Corporation, Austin, TX). Cytokine levels (pg/mL) were automatically calculated from standard curves using Bio-Plex Manager software (v. 6.1, Bio-Rad).

Biological sample location was stratified across 5 assay plates such that all trial arms had approximately equal representation on each plate. All samples from a given participant were grouped onto the same plate to reduce batch effects. Location within the plate was randomized to reduce location-based bias. For each biological sample, the concentrations of all cytokines listed above were measured in two technical replicates. The average was used for downstream data analysis. Average values falling below the assay range were reassigned to 1/2 of the lower bound of the assay range.

